# Automated ELISA on-chip for the detection of anti-SARS-CoV-2 antibodies

**DOI:** 10.1101/2021.08.05.21261664

**Authors:** Everardo González-González, Ricardo Garcia-Ramirez, Gladys Guadalupe Díaz-Armas, Miguel Esparza, Carlos Aguilar-Avelar, Elda A. Flores-Contreras, Irám Pablo Rodríguez-Sánchez, Jesús Rolando Delgado-Balderas, Brenda Soto-García, Diana Aráiz-Hernández, Marisol Abarca-Blanco, José R.Yee-de León, Liza P. Velarde-Calvillo, Alejandro Abarca-Blanco, Juan F. Yee-de León

## Abstract

The COVID-19 pandemic has been the most critical public health issue in modern his-tory due to its highly infectious and deathly potential; and the limited access to massive, low-cost, and reliable testing has significantly worsened the crisis. The recovery and the vaccination of millions of people against COVID-19, have made serological tests highly relevant to identify the presence and levels of SARS-CoV-2 antibodies. Due to its advantages, microfluidic-based technologies represent an attractive alternative to the conventional testing methodologies used for these purposes. In this work, we describe the development of an automated ELISA on-chip capable of detecting anti-SARS-CoV-2 antibodies in serum samples from COVID-19 patients and vaccinated individuals. The colorimetric reactions were analyzed with a microplate reader. No statistically significant differences were observed when comparing the results of our automated ELISA on-chip against the ones obtained from a traditional ELISA on a microplate. Moreover, we demonstrated that it is possible to carry out the analysis of the colorimetric reaction by performing basic image analysis of photos taken with a smartphone, which constitutes a useful alternative when lacking specialized equipment or a laboratory setting. Our automated ELISA on-chip has the potential to be used in a clinical setting and mitigate some of the burden caused by testing deficiencies.

## 1. Introduction

The coronavirus disease 2019 (COVID-19) is caused by the severe acute respiratory syndrome coronavirus 2 (SARS-CoV-2) and was officially declared as a pandemic by the World Health Organization (WHO) in March 2020. By July 2021, public records registered over 200 million infections, 4 million deaths, and 3,000 million COVID-19 vaccine doses administered worldwide; additionally, several genetic variants have been identified so far [1–3]. Since its inception, the attempts to contain this disease have consisted in confinement and molecular diagnostics. The real-time reverse transcription-polymerase chain reaction (RT-qPCR) is considered the gold standard method to diagnose COVID-19, which is based on the amplification of SARS-CoV-2 genes (*N, E, RdRp, orf1a*, and o*rf1b*) and its detection by fluorescent reporters from nasopharyngeal swab samples [4,5]. Other molecular methods such as immunoassays, directed to detect viral antigens or an-ti-SARS-CoV-2 antibodies, have proven to be of utmost importance for the pandemic mitigation [6–8]. Understanding how the levels of anti-SARS-CoV-2 antibodies fluctuate in recovered COVID-19 patients and vaccinated individuals is fundamental to better understand the disease, especially when discrepancies have been found on previous reports [9–12]. Under the current circumstances, where millions of viral infections exist, is un-deniable that vaccination is of vital importance, and immunoassays have taken a particularly relevant role in identifying and monitoring patients’ immune responses over time. Therefore, several research groups have proposed various COVID-19-related immunoassays which are mainly based on viral antigens, such as the spike protein [13], receptor-binding domain (RBD) [14], and nucleoprotein [15] and include different strategies ranging from the traditional enzyme-linked immunoassay (ELISA) to more complex microfluidic immunoassays [16–18].

Through the course of the COVID-19 pandemic, several diagnostic challenges have emerged, particularly regarding the millions of tests require to face the disease spreading. Microfluidic technologies are a promising approach that could solve some of those problems, because they enable the integration and automation of complete diagnostic protocols in a single chip (i.e., lab-on-a-chip); including all the steps ranging from sample preparation to the detection and quantification of the analyte of interest [19]. A lab-on-chip meets the requirements of a traditional laboratory setting but with the advantages of miniaturization, reducing the volume of reagents and incubations times [20]. Some of the difficulties that have restricted the widespread use of these technologies in medical and biomedical applications are the manufacturing processes to mass-produce these micro-fluidic devices and its subsequent functionalization to attach biomolecules to their surface [21,22].

To address some of the diagnostic challenges, derived from testing deficiencies, we developed an automated ELISA on-chip, capable of detecting anti-SARS-CoV-2 antibodies in serum samples from COVID-19 patients and vaccinated individuals. Most microfluidic immunoassays use polydimethylsiloxane (PDMS) or polymethyl methacrylate (PMMA) microfluidic devices [23–25], that carry inherent disadvantages regarding the complexity of its functionalization and its compatibility with large-scale manufacturing processes. However, in our work we utilized a microfluidic device made of polystyrene (PS), which is an ideal material for this application, due to its mass manufactured feasibility and its hydrophobic properties, making it unnecessary to add any extra treatment to attach biomolecules to its surface [26]. Commercially available microfluidic instrumentation was used to automate every step of the on-chip immunoassay from antigen immobilization to the detection of anti-SARS-CoV-2 antibodies. In order to assess the clinical potential of our automated ELISA on-chip we compared the results against the ones obtained with a traditional ELISA, made on a microplate, and we found no statistically significant differences between both immunoassay formats. With a minimum sample manipulation, a reduced volume of reagents, and being time effective, the use of microfluidic technologies becomes a valuable alternative for mass testing, especially in a pandemic setting.

## 2. Materials and Methods

### 2.1 Sample collection and preparation

Blood samples were collected according to the protocol approved by the Institutional Review Board of the Alfa Medical Center with number AMCCI-TECCOVID-001. All the participants provided written informed consent before sample collection, which took place in a clinical laboratory, that followed the guidelines established by Official Mexican Standards: NOM-007-SSA3-2011, NOM-087-SEMARNAT-SSA1-2002, NOM-010-SSA2-2010, NOM-006-SSA2-2013, and NMX-EC-15189 IMNC-2015. Blood samples were centrifuged at 1,000 g for 10 minutes at 4°C to separate the serum. In this work, a total of 22 serum samples were analyzed, 7 of them belonged to COVID-19 patients (samples from 3- and 7- months post-infection were available for 2 patients, while only samples from 7-months post-infection were available for the other 5 patients), 4 were from vaccinated volunteers (samples from 0- and 60-days post-vaccine were available for 2 patients, while only samples from 60-days post-vaccine were available for the other 2 patients), and 7 corresponded to healthy volunteers (2 samples were taken before the COVID-19 pandemic started). BSA was included as a negative control. All positive and negative samples were confirmed by qRT-PCR. All procedures involving human participants were performed in accordance with the 1964 Helsinki declaration and its later amendments or comparable ethical standards.

### 2.2 Traditional ELISA on a microplate

A traditional ELISA was performed using a 96-well microplate (Corning Inc., MA, USA) to compare those results against the ones obtained with our automated ELISA on-chip. Firstly, 100 µL of a PBS suspension containing 1 µg/mL of the complete spike protein (Sino Biological Inc., PA, USA) were deposited in each well, followed by a 1 hour incubation at room temperature. Afterward, three washes were made using a wash buffer (WB = PBS containing 0.05% TweenTM 20 (Thermo Fisher Scientific, MA, USA)). Blocking was made by incubating for 1 hour 200 µL of 5% skim milk (Sigma-Aldrich, MA, USA) at room temperature. Subsequently, another round of three washes were carried out using the WB. Then, the serum samples (1:100 dilution) were added to the microplate and incubated for 1 hour at room temperature, to later be washed three times with WB. Next, a 1 hour incubation of 100 µL of anti-human IgG conjugated with HRP (1:15,000 dilution) (Thermo Fisher Scientific, MA, USA) was performed, at room temperature, to identify the presence of anti-spike antibodies; followed by three washes with WB. Finally, 100 μL of 1-StepTM Ultra TMB-ELISA (Pierce Biotechnology Inc., IL, USA) were used to reveal the reaction, and the reaction was stopped by adding 100 μL of 1M H2SO4.

### 2.3 Assay’s methodology and experimental setup of the automated ELISA on-chip

The methodology followed in the automated ELISA on-chip assay and a diagram that illustrates the experimental setup implemented are displayed in Figures 1A and 1B, respectively. The reagents used and the conditions at which these were passed through the microfluidic device are specified in Table 1.

**Table 1.** Established protocol for our automated ELISA on-chip

| Step | Reagent | On-chip flow dynamics |
| --- | --- | --- |
| **1. Antigen immobilization** | 1 μg/mL spike protein in PBS | 500 μL/min (30 s) - incubation (1 hr/RT) |
| **2. Wash** | 0.05% Tween-20<sup>TM</sup> in PBS | 500 μL/min (30 s) |
| **3. Blocking** | 5% skim milk in PBS | 500 μL/min (1 min) - incubation (30 min/RT) |
| **4. Wash** | 0.05% Tween-20<sup>TM</sup> in PBS | 500 μL/min (1 min) - 3 times |
| **5. Pumping of samples** | Serum (diluted in PBS 1:100) | 500 μL/min (1 min) - incubation (50 min/RT) |
| **6. Wash** | 0.05% Tween-20<sup>TM</sup> in PBS | 500 μL/min (1 min) - 3 times |
| **7. anti-IgG-HRP binding** | anti-IgG-HRP (diluted in PBS 1:15,000) | 500 μL/min (30 s) - incubation (50 min/RT) |
| **8. Wash** | 0.05% Tween-20<sup>TM</sup> in PBS | 500 μL/min (1 min) - 3 times |
| 9. HRP reaction | TMB-ELISA | 50 μL - incubation (3 min/RT) |
| 10. Reaction halt | 1M H<sub>2</sub>SO<sub>4</sub> | 50 μL (final reaction) |

**Figure 1.**
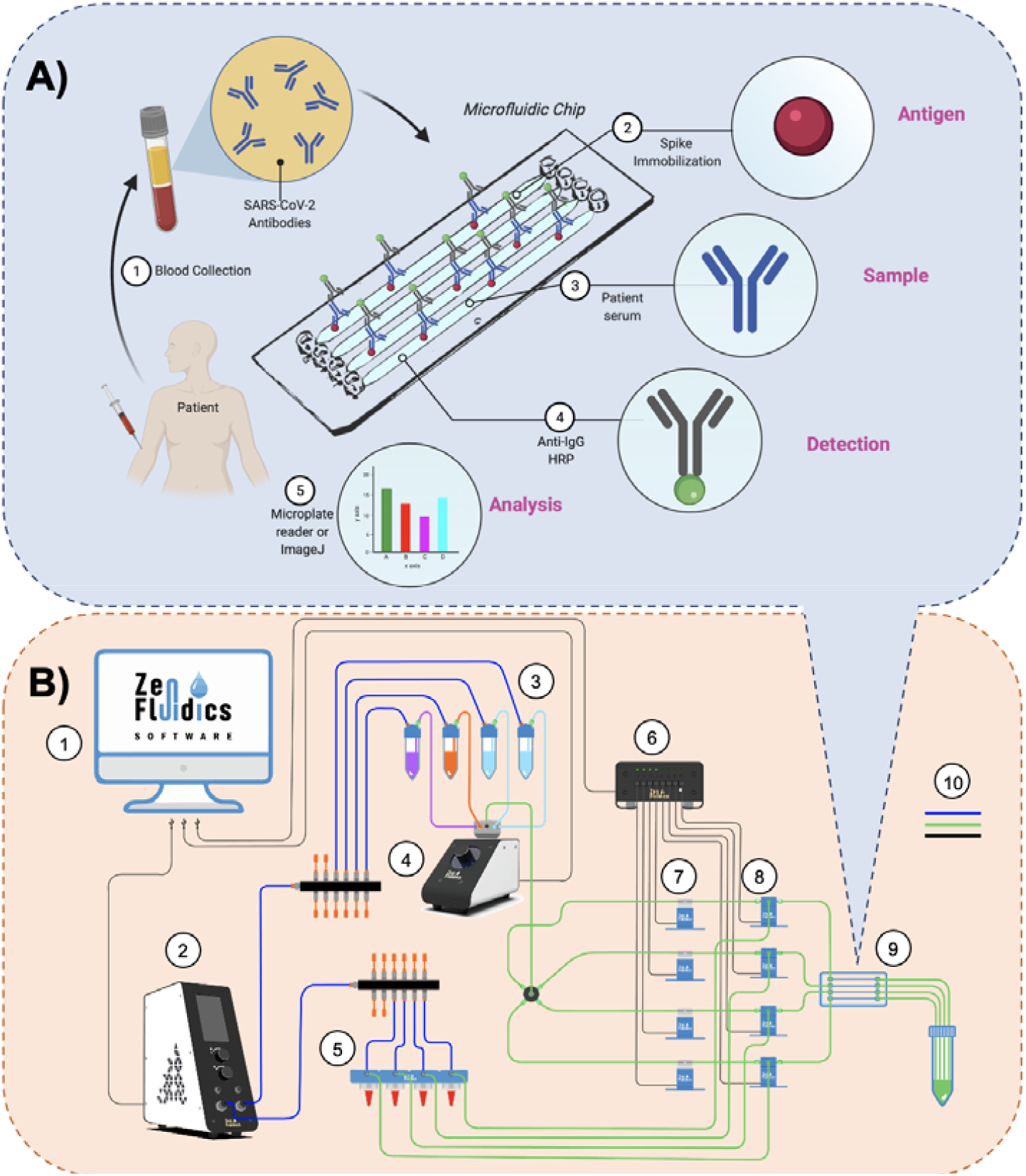
Assay’s methodology and experimental setup. A) The methodology followed in our automated ELISA on-chip assay included the steps listed below: 1) Blood collection and serum extraction, 2) On-chip immobilization of viral antigens (SARS-CoV-2 spike protein), 3) Pumping serum samples through the microfluidic device (if present, anti-SARS-CoV-2 antibodies interact with the immobilized SARS-CoV-2 spike proteins), 4) Pumping anti-IgG-HRP through the micro-fluidic device to detect IgG antibodies, and 5) Analysis of the colorimetric reaction, either by re-covering the resulting reaction and performing the analysis with a microplate reader or by car-rying out a color intensity analysis using ImageJ software. B) Diagram of the experimental setup assembled for our automated ELISA on-chip. The components are listed below: 1) Software that controls all the peripheral devices of the setup, 2) Flow control unit, 3) ELISA reagents (wash buffer, blocking buffer, and anti-IgG-HRP conjugate antibody), 4) 12/1 bidirectional microfluidic rotary valve, 5) Serum samples, 6) Microfluidic valve controller, 7) Pinch valves, 8) 3/2-way switching valves, 9) Microfluidic device, and 10) Connections between components and reser-voirs (color code: blue is for pneumatic connections, green is for fluid connections, and black is for electrical connections).

Commercially available microfluidic instrumentation was used to assemble the experimental setup that enabled the automation of our ELISA on-chip assay, from antigen immobilization to the detection of an-ti-SARS-CoV-2 antibodies. A PS microfluidic device with four straight channels (50 µL volume capacity/channel) (microfluidic ChipShop, Jena, Germany), a flow control unit (Zen Fluidics, TX, USA), a 12/1 bidirectional microfluidic rotary valve (Zen Fluidics, TX, USA), a microfluidic valve controller (Zen Fluidics, TX, USA), a set of 4 pinch valves (Zen Fluidics, TX, USA), and a set of 4 3/2-way switching valves (Zen Fluidics, TX, USA) were the main components that constitute this setup. All protocol steps were programmed using the Zen Lab software (Zen Fluidics, TX, USA), which also served to control all the components of the setup. All the pneumatic, fluidic, and electrical connections are also specified in the diagram depicted in Figure 1B. Additionally, a photograph of the experimental setup is shown in Figure 2.

**Figure 2.**
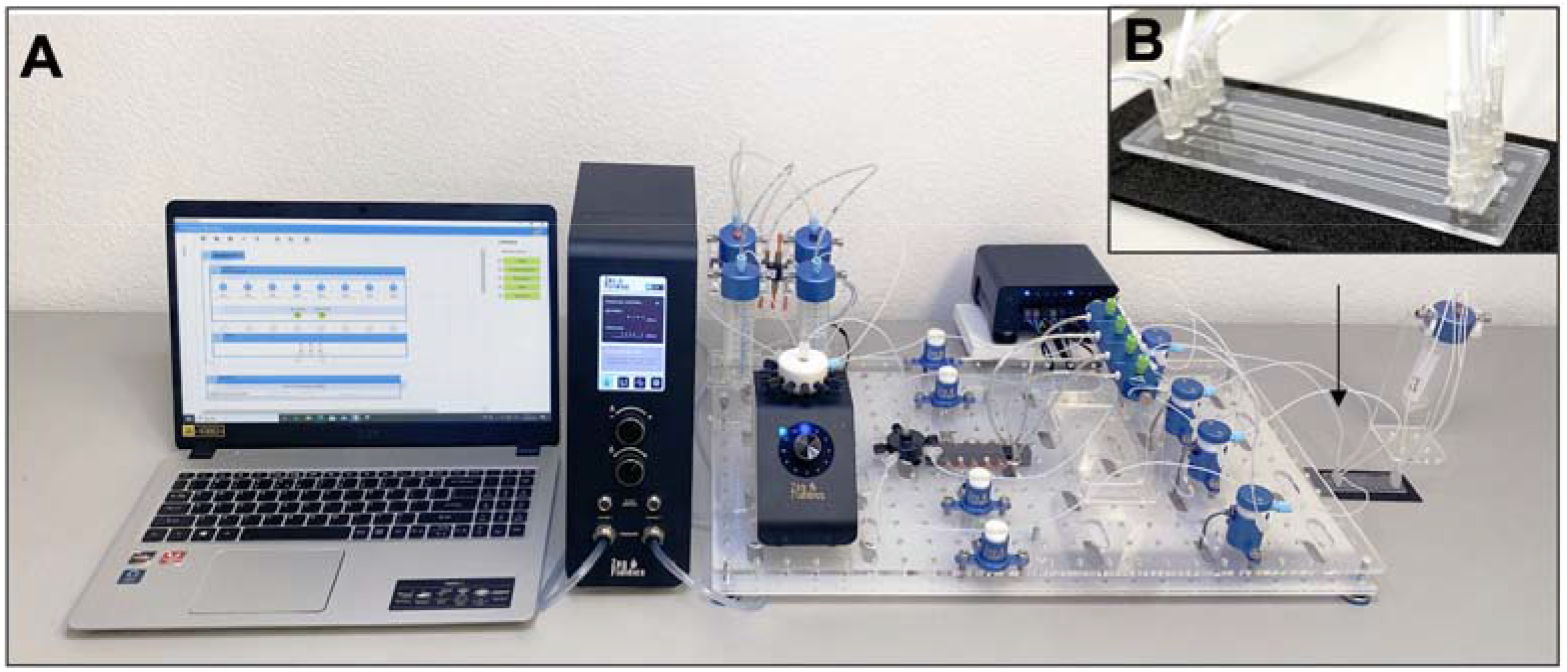
A) Photograph of the experimental setup of our automated ELISA on-chip. B) Picture of the microfluidic device used in our setup.

### 2.4 Data analysis

The colorimetric reactions obtained when performing the traditional ELISA and our automated ELISA on-chip were analyzed with two different methods. The first one used a microplate reader (BioTek Instruments, VT, USA) and measured the reaction at 450 nm. The second approach was based on taking photos with a commercial smartphone, of the microplate and microfluidic devices, under same lighting conditions, for further color intensity analysis using the color histogram plugin of the ImageJ software (NIH, MD, USA) to assign a value to each reaction.

### 2.5 Statistical analysis

The Kolmogorov-Smirnov test was used to determine if our data followed a normal distribution. The results showed non-parametric behavior; therefore, the statistical analysis was performed with the Wilcoxon test, using a p-value < 0.01 and the IBM SPSS Statistics 27 software (IBM Corporation, NY, USA). All samples were run in triplicate. DataGraph version 4.7 (Visual Data Tools Inc., NC, USA) and Microsoft Excel (Microsoft Corporation, WA, USA) were employed to graph the results.

## 3. Results and Discussion

### 3.1. Immunoassays’ comparison

To validate our results, we performed a comparison between the traditional ELISA and our automated ELISA on-chip, in which anti-SARS-CoV-2 antibodies were detected. For both immunoassay formats, the colorimetric reactions derived from the horseradish peroxidase were analyzed with a microplate reader (Figure 3A). The 22 samples used in this work were collected from COVID-19 patients, vaccinated individuals, and healthy volunteers. The results showed a non-parametric distribution; therefore, the Wilcoxon test was employed to make the statistical analysis, indicating that there wasn’t a statistically significant difference between both immunoassays.

**Figure 3.**
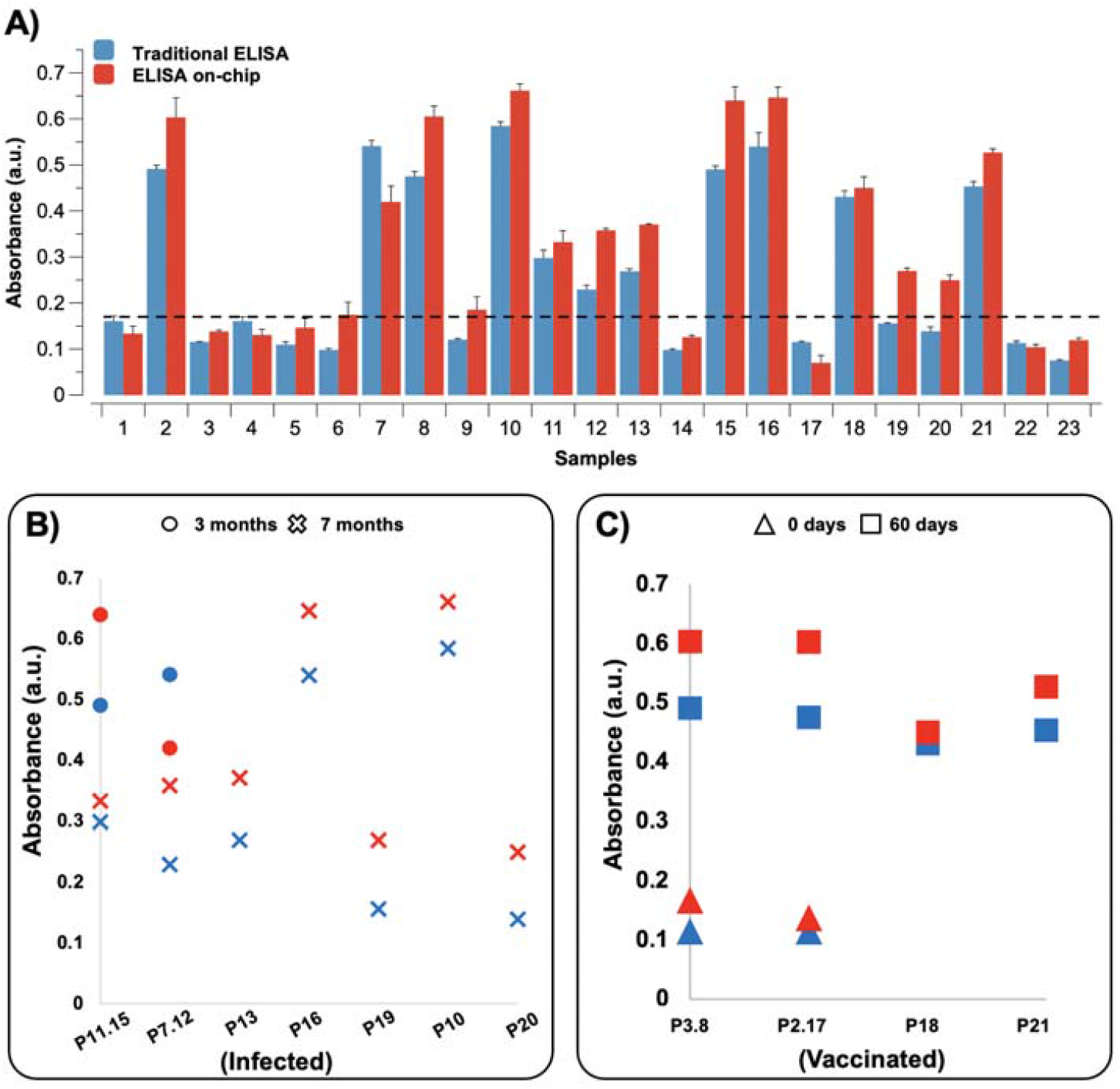
Absorbance comparison between the traditional ELISA and our automated ELISA on-chip. The resulting colorimetric reactions from both immunoassay formats were analyzed with a microplate reader. A) Measured absorbance of the 22 serum samples and 1 negative control. The dotted line indicates the average absorbance measured in serum samples from healthy subjects. B) Measured absorbance of the serum samples collected from COVID-19 patients 3 months (circle) and 7 months (cross) after infection. C) Measured absorbance of the serum samples collected from vaccinated individuals prior being vaccinated (triangle) and 60 days after vaccination (square).

### 3.2. Detection of anti-SARS-CoV-2 antibodies post-infection

We also monitored the presence of anti-SARS-CoV-2 antibodies in 2 patients diagnosed with COVID-19 at 3- and 7-months post-infection. In these patients, a decrease in absorbance was observed in samples collected 7 months after infection compared to the samples collected 3 months post-infection.

Furthermore, for the other 5 patients, only samples from 7 months post-infection were available, which showed comparable, or even higher antibody levels. The data obtained from both immunoassay formats, the traditional ELISA, and the automated ELISA on-chip, are displayed in Figure 3B.

The immune response after COVID-19 infection is heterogeneous among subjects, whereas, for diseases caused by other coronaviruses, such as SARS-CoV and MERS-CoV, antibodies can be detected up to 34 weeks after being infected [27,28]. As an example, Labriola et al. reported a decline in anti-spike and anti-nucleocapsid IgG levels in patients that tested positive for COVID-19, 3 months after infection, while Dan et al. reported stable levels of anti-SARS-CoV-2 antibodies after 8 months from viral infection. Despite this heterogeneity, the immune response seen in patients after infection agrees with what has been described in other reports [29–32].

### 3.3. Detection of anti-SARS-CoV-2 antibodies post-vaccine

In this study, we also analyzed serum samples from four vaccinated individuals. Blood was collected and processed 60 days after vaccination. To assess the increase of anti-SARS-CoV-2 antibodies after inoculation, we collected two samples from two of those individuals before they were vaccinated. As seen in Figure 3C, using both assay formats, high levels of anti-SARS-CoV-2 antibodies were detected in the four samples extracted 60 days after vaccination. Furthermore, the immune response triggered by the administration of this biologic can be observed on the two patients that had samples available before and after vaccination.

In an effort to control this pandemic, different immunization strategies have been developed. Currently, the FDA has approved three different vaccines based on either adenovirus or lipidic complexes containing mRNA molecules. In a recent work by Jalkanen et al., an increase in the production of anti-SARS-CoV-2 IgG antibodies was observed at six weeks after the administration of the mRNA vaccine (BNT162b2). Additionally, Stephenson et al. reported a detectable increase of anti-SARS-CoV-2 antibodies eight days after the administration of the adenoviral Ad26.COV2.S vaccine, detecting higher concentrations of these antibodies 57 and 71 days after inoculation. The results obtained in this work are in line with the results reported in other studies where these biologics were assessed, which showed an increase of anti-SARS-CoV-2 antibodies after the application of a vaccine [33–35].

### 3.4 Image analysis of colorimetric reactions

Diagnostic tests usually require expensive equipment and/or specialized technicians, which complicates its implementation in underdeveloped communities. Therefore, we propose to use photographs taken with a commercial smartphone to perform the analysis of the resulting colorimetric reaction as a simple and low-cost alternative to microplate readers. We were able to detect anti-SARS-CoV-2 antibodies in serum samples from COVID-19 patients, and individuals vaccinated using both immunoassay formats and assigning a value to each reaction by analyzing the color intensity of every picture taken. Figure 4A visually describes the analysis process, while pictures of the resulting reactions when performing the traditional ELISA and our automated ELISA on-chip are shown in Figure 4B. Since both results, the ones obtained from the microplate reader and the analysis with ImageJ, showed a non-parametric distribution, the Wilcoxon test was used to make the statistical analysis, revealing that there were no statistically significant differences between these two approaches. These results are displayed in Figure 4C. There is a clear trend towards the development of point of care (POC) diagnostic devices to mitigate healthcare deficiencies and improve the health conditions of people living in underserved communities. Its popularization is largely due to their compatibility to be mass produced, ease of operation, small footprint, lower costs, and the capacity to rapidly generate accurate and reliable results. To date, numerous POC devices have been developed for its use in a broad range of medical and biomedical applications, including the detection of infectious diseases, such as Zika [36], Ebola [37], and now, COVID-19 [38]. Furthermore, various have been cleared by FDA and are commercially available [39]. There are currently many things left to know regarding the immune response to COVID-19 and its vaccines. To facilitate this knowledge, it is extremely necessary a re-liable, high-throughput, and automated method for quantitatively measure an-ti-SARS-CoV-2 antibodies. Therefore, in this manuscript, we propose an automated ELISA on-chip to detect anti-SARS-CoV-2 antibodies in serum samples. This approach can still increase the number of samples to be processed as well as reducing the time needed to do so; it could also be used for other applications.

**Figure 4.**
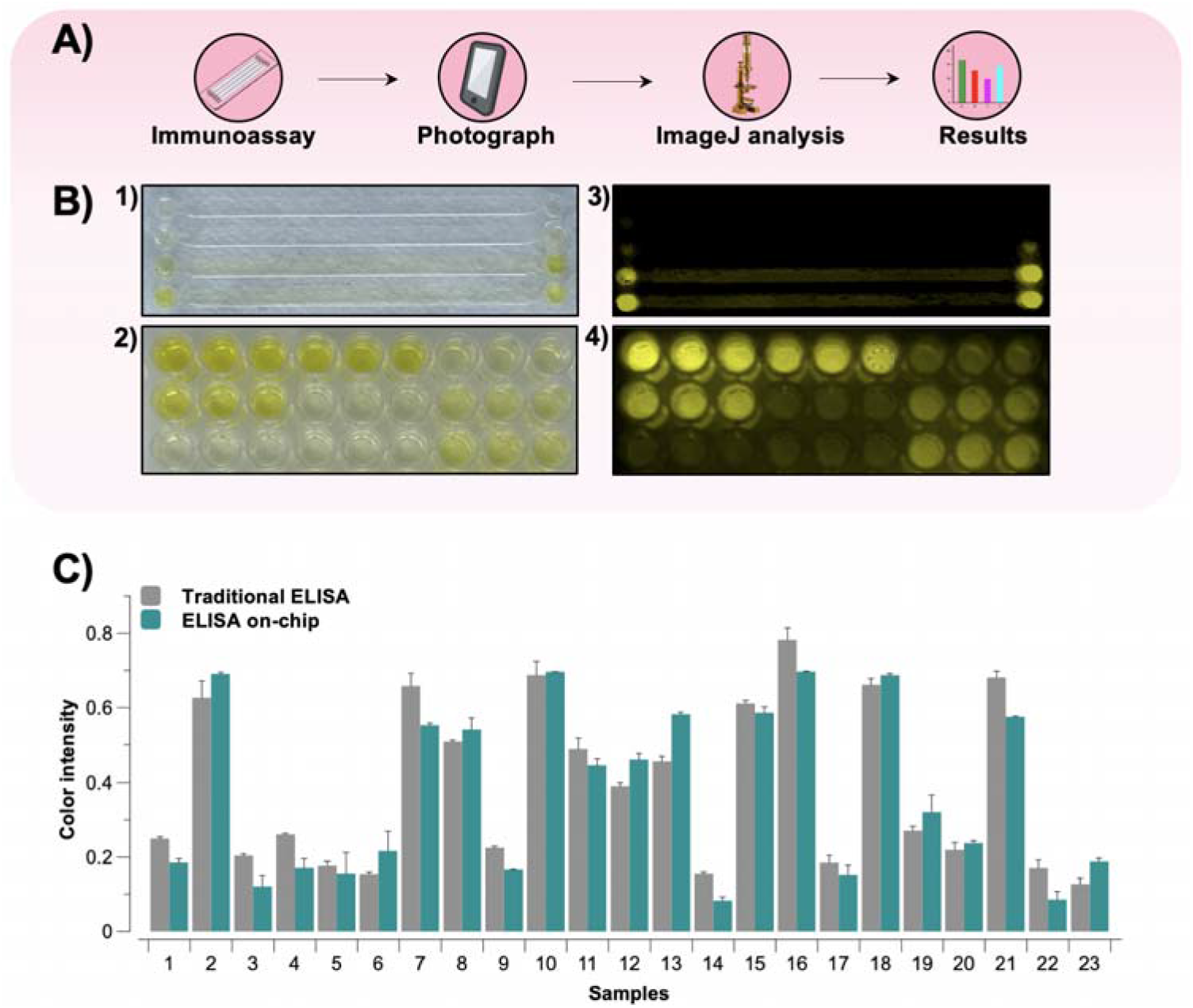
A) Analysis methodology: First, the immunoassay is performed. Then, a photograph of the substrate, either a microplate or a microfluidic device, is acquired with a smartphone, fol-lowed by a color intensity analysis of the resulting colorimetric reactions using the ImageJ soft-ware or any other similar software. Finally, the results are plotted and compared against the ones obtained from other samples. B) Photographs of the resulting reactions when performing (1) our automated ELISA on-chip and (2) the traditional ELISA, and its corresponding processed images (3, 4). C) Comparison of the results obtained by image analysis of the colorimetric reactions with both immunoassay formats from the 22 serum samples and 1 negative control.

## 5. Conclusions

Microfluidic technologies can enable the integration and automation of complete diagnostic protocols in a single chip, making them a truly advantageous option for mass testing, particularly in a pandemic setting. This manuscript presented a novel format for the detection of anti-SARS-CoV-2 antibodies by using an automated immunoassay on-chip, while comparing it to the gold standard methodology utilized for immunoassays. Our work revealed that there were no significant differences between the results of both immunoassays, which were obtained from: 1) A microplate reader, the most common instrument to measure absorbance in immunoassays, and 2) An image processing software that analyzed a picture taken with a smartphone. Besides being reliable, this last method constitutes a fast, easy, low-cost, and extremely valuable alternative for analyzing colorimetric assays, especially when a microplate reader or a specialized technicians are not available to underserved communities.

## Data Availability

We reported the results in this manuscript

## Author Contributions

Conceptualization, E.G.-G. and J.F.Y.-d.L.; methodology, E.G.-G., R.G.-R., G.G.D.-A., and J.R.D.-B.; software, J.R.Y.-d.L., C.A.-A., and M.E.; validation, B.S.-G., D.A.-H., G.G.D.-A, and R.G.-R.; formal analysis, E.A.F.-C. and J.F.Y.-d.L.; investigation, J.F.Y.-d.L. and E.G.-G.; resources, M.A.-B. and I.P.R.-S.; data curation, E.A.F.-C. and E.G.-G.; writing—original draft preparation, E.G.-G. and J.F.Y.-d.L.; writing—review and editing, E.G.-G., C.A.-A., G.G.D.-A., R.G.-R., and J.F.Y.-d.L.; visualization, J.R.Y.-d.L., C.A.-A., and M.E.; supervision, A.A.-B., L.P.V.-C., and J.F.Y.-d.L.; project administration, A.A.-B., L.P.V.-C., and J.F.Y.-d.L.; funding acquisition, A.A.-B., L.P.V.-C., and J.F.Y.-d.L. All authors have read and agreed to the published version of the manuscript. Funding: This research received no external funding.

Institutional Review Board Statement: Blood samples were collected according to the protocol approved by the Institutional Review Board of the Alfa Medical Center with number AMCCI-TECCOVID-001. All procedures involving human participants were performed in ac-cordance with the 1964 Helsinki declaration and its later amendments or comparable ethical standards.

Informed Consent Statement: Informed consent was obtained from all subjects involved in the study. Acknowledgments: We acknowledge the support in figure design by Alitzel Trueba Zúñiga.

